# Pathogenic ultra-rare variants in *SLC6A1*, *SLC6A11*, *GAD1* and *GAD2* are new & recurrent GABAergic loci for GGE syndromes

**DOI:** 10.1101/2025.04.01.24316792

**Authors:** Seo-Kyung Chung, Kate V. Everett, Rhys H. Thomas, Ed Dudley, Adam T. Higgins, Charlotte A. Jones, Anna V. Derrick, Jeff S. Davies, William O. Pickrell, Susannah T. Bellows, Robert J. Harvey, Peter S. Bergin, Jonathan G.L. Mullins, Ingrid E. Scheffer, Samuel F. Berkovic, Mark I. Rees

## Abstract

**Purpose:** There is a wealth of biological evidence that supports the concept that GABAergic inhibition is causal to some genetic generalized epilepsy (GGE) syndromes. Much is known about postsynaptic GABA_A_R channelopathies, however, the presynaptic determinants are less well-defined such as the role of GABA transporters (*SLC6A1* / GAT1; *SLC6A11 /* GAT3), and GABA biosynthesis biology (*GAD1* / GAD67; *GAD2* / GAD65).

**Methods:** In response, our study screened four presynaptic GABAergic genes in an international cohort of 708 adults and children with epilepsy, predominantly with GGE syndromes, diagnosed within tertiary epilepsy centers.

**Results:** We identified 15 novel / ultra-rare heterozygous *SLC6A1*, *SLC6A11, GAD2, GAD1* variants in 31 unrelated people with epilepsy, of whom 28 had GGE syndromes. A range of missense, nonsense and splice-site variants were discovered in addition to one digenic case with *SLC6A1* & *GAD2* variants. *SLC6A1* genotypes identified in this study show reduced GAT1 activity, quantitatively linked to epilepsy severity and intellectual disability (ID). Functionally validated *SLC6A11* and *GAD2* variants are a novel GGE finding, where reduced activity of GAT3 uptake or GAD65 enzyme activity is linked to epilepsy severity. Recessive *GAD1* loss-of-function variants have recently been described as the cause of developmental and epileptic encephalopathy cases. Here we present the first two gain-of-function *GAD1* variants as a novel GAD67 mechanism in epileptogenesis.

**Conclusion:** This further contributes to the association of GABA proteome with GGE and links subtle quantal GABA sensitivities to the genesis of cortico-thalamic absence seizures in GGE syndromes.

## Introduction

Genetic generalized epilepsy (GGE; MIM: 600669) comprises a number of well delineated electro-clinical syndromes differentiated by seizure type, age of onset and EEG features.^1^ There is a substantive genetic component to their etiology including copy number variation, rare and common variants.^2–4^ Mutations in postsynaptic GABA_A_ receptor subunits linked to GGE affect the inhibition of excitatory cells in the mammalian brain.^5–9^ However, most of the genetic contribution to these ‘genetic epilepsies’ remain undiscovered. Meanwhile, there are multiple neurological conditions linked to cognate synaptopathies that involve both postsynaptic receptors and presynaptic transporters.^10–11^

Our *a priori* hypothesis was that the GGEs hold an excess of variants in both pre- and post-synaptic GABAergic genes. We restricted the search to four genes: specifically, *SLC6A1, SLC6A11, GAD1, GAD2*. The genes *SLC6A1* and *SLC6A11* encode the presynaptic Na^+^/Cl^-^ co-transporters GABA Transporter-1 (GAT1) and GABA Transporter-3 (GAT3) respectively and they ensure synaptic reuptake of vesicular-released GABA from the presynaptic cleft or extrasynaptic reuptake into astroglial cells.^12–18^ Generally, GAT1 is located on presynaptic sites whilst GAT3 is preferentially localized on astroglial cells and recycles excess GABA that has migrated into the neuronal periphery. In the thalamus, GAT3 is more abundant on neurons, indicating a unique thalamocortical role, sensitivity, and functional specificity.^19–20^ *GAD1* and *GAD2* encode the main biosynthetic enzymes, GAD67 and GAD65, that collectively produce presynaptic vesicular GABA in neurons.^21^ GAD65 is the reactive on/off biosynthesis isoform, which can be rapidly activated and deactivated upon pyridoxal 5′-phosphate (PLP) binding. GAD65 antibodies are associated with a range of neurological disorders including limbic encephalitis, stiff person syndrome and epilepsy.^22,23^ GAD67, in contrast, is responsible for steady-stream constitutive production of neuronal GABA and is essential for normal neuronal function and development.

During our study, we and other colleagues reported heterozygous loss-of-function variants in *SCL6A1* in myoclonic astatic epilepsy (MAE) and absence seizures with intellectual disability.^24–30^ More recently, bi-allelic and homozygous *GAD1* mutations have been linked to a severe epileptic encephalopathy (EE) supporting our rationale for examining these genes in GGE.^26, 31–32^

## Materials and Methods

### Cases and Participants

708 people with epilepsy, predominantly with GGE, were ascertained from academic epilepsy centres (**Supplementary Table 1**). Each case was diagnosed and ethically consented into studies by specialist epileptologists in tertiary healthcare settings. Case ascertainment was frequently accompanied by a full medical record review and therefore longitudinal syndrome transition has been captured, such as Childhood Absence Epilepsy (CAE) transitioning to Juvenile Myoclonic Epilepsy (JME). Syndrome classification was based on notes review, rather than a proforma, and so all evidence was available to review.

### Genetic Screening and Bioinformatics

The entire coding regions of the four GABAergic genes were analysed using high-throughput screening process and Sanger sequencing. Population studies of potential novel (not in the population) and very-rare variants (Mean Allele Frequency, MAF<0.01, study criteria) were sourced from dbSNP, and population-stratified gnomAD data (https://gnomad. broadinstitute.org). Bioinformatic databases were scrutinised (SIFT, Polyphen-2, Grantham scores) to indicate protein-damaging consequences or establish phylogenetic conservation.

### Structural Protein Modelling

The structural modelling of GAT1 and its variants exploited the recently determined cryo-EM structure, Protein Data Bank (PDB):7Y7W.^33^ GAT3 WT and variants were modelled from the basis of the relevant AlphaFold structure (for UniProt P48066).^34^ Complete structures for GAD65 and GAD67 are not available. For these proteins, modelling of wild-type and gene-variants was carried out as described previously^35–36^ using an established homology modelling pipeline built with the open source Biskit structural bioinformatics platform, which scans the entire PDB for candidate homologies.^37^ The protein modelling platform is capable of modelling both wild-type structure and conformational differences of variant forms.^38^ The best pipeline homology attained for GAD65, was 99%, with a portion of the X-Ray structure of the 65kDa isoform GAD65 (PDB: 2OKK); and for GAD67, 99% with X-Ray structures of the 67kDa isoform GAD67 (PDB: 2OKJ and 3VP6). The compound structures for GABA and Glutamic Acid were obtained from PubChem and docked by superposition upon the positions of the appropriate bound ligands in the above crystal structures.^39^ All models were visualized and analysed using the graphics program Chimera.^40^

### Generation of Expression Constructs

The C-terminally DDK-tagged wild-type gene-constructs were externally sourced for all four GABA genes and sequence verified (OriGene Technologies; pCMV6-entry). The 4.9Kb Kanamycin resistant vector contained the human CMV promotor, an ideal Kozak sequence prior to the insert, and a C-terminal DDK tag sequence. A stop codon was introduced to all WT constructs to remove the DDK tags representing native proteins at physiological conditions. The GABAergic variants (**Supplementary Table 2, 3 and 4**) were introduced by mutagenesis of wild-type constructs, purified using a maxi-prep kit (Qiagen) and the entire coding regions were sequence validated. The novel 1bp deletion variant, c.1780delA, of *SLC6A1* is expected to create a frameshift p.(Ser594Alafs*22) outcome, adding 49 extra nucleotides to the *SLC6A1* coding region. Insertion of the additional base pairs was prepared using fusion of complementary sequences (Megaprimer PCR) by targeting a defined DNA fragment into a precise region of the WT GAT1 construct followed by a mutagenesis PCR. The expression of all constructs was validated using cultured HEK293 cells and Western blot analysis.

### Cell-Surface Expression

Biotinylation assay was undertaken on all GAT1 and GAT3 variants for any aberrant trafficking outcomes as previously outlined in Chung *et al* 2013.^41^ 24 hrs after transfection, surface expression in HEK293 cells were investigated using a cell membrane-impermeable reagent Sulfo-NHS-LC-Biotin (Pierce Biotechnology).

### A Mass Spectrometry GABA Transporter Assay

This was developed for GAT1 and GAT3 variant analysis using an isotopically labelled GABA (CDN Isotopes) and Gas-Chromatography electron-impact Mass Spectrometry (GCMS). All GAT1 and GAT3 variants were tested with the GABA transporter assays using modified methods of previously published study.^42^ Dose-response curves for the GCMS assay were obtained for WT GAT1 (K_m_=13.07 ± 2.291µM) and GAT3 (K_m_=18.31 ± 2.728µM) and are comparable to previous platforms (**Supplementary Fig. 1**). HEK293 cells were seeded on 12 well plates and 24 hours later, GAT1 and GAT3 WT and variants were transfected using TurboFectin 8.0 (Origene). 24 hours post-transfection, cells were washed with uptake buffer including 1M 4- (2-hydroxyethyl)-1-piperazineethanesulfonic acid (HEPES, Gibco) in HBSS solution then incubated with the uptake buffer for 25 min at 37°C. Following addition of various concentrations of GABA (0.00-0.5mM), cells were incubated for 6 min at 37°C. Uptake was terminated by adding cold wash buffer (154mM of Ammonium aetate) and cells were lysed by incubating for 2 hrs at 37°C with lysing buffer containing 87.5% of Acetonitrile. Lysed samples were vacuum dried then tested using GCMS at the EPSRC National Mass Spectrometry Unit in Swansea. Dried samples were reconstituted in 30µl of pyridine containing 15mg/ml methoxylamine hydrochloride and incubated at 70⁰C for 1 hour before adding 50µl MSTFA (Thermofinnigan, UK) and heating at 40⁰C for 90 minutes. 1µl of the sample was injected onto a 6890 GC (Agilent technologies, UK) in splitless mode, an injector temperature of 250⁰C and a helium gas flow of 1ml/minute.

The metabolites extracted and derivatised were separated on a Thermo TR-5 ms SQC GC column (30 m × 0.25 mm × 0.25μm) (Thermo Scientific, UK) using a temperature gradient from 60 to 180⁰C over 12 minutes and an increase of 4⁰C per minute to a final temperature of 300⁰C. The GC eluent was analysed using a 5975 inert mass selective detector (Agilent technologies, UK) in electron ionisation mode, scanning from m/z 50-650 with a source temperature of 230⁰C and a quadrupole temperature of 150⁰C. The deuterated GABA was detected via the prominent ion at m/z 306, with a second ion at m/z 174 being used to confirm the identification. An extracted ion chromatogram of m/z 306 was used to determine the peak area for the GABA m/z 306 ion and this was normalised to the total ion chromatogram for each sample.

### An enhanced GABase GAD Enzymatic Assay

Using a fluorescence-based microplate method, we employed an enhanced GAD enzymatic assay to quantify the amount of GABA synthesized by WT GAD and variant GAD enzymes. GABase (Sigma-Aldrich) contains two enzymes (Gamma-aminobutyric acid aminotransferase and succinic semialdehyde dehydrogenase) and is classically used to measure intra and extracellular level of GABA by coupling with formation of NADPH which absorbs at 340nm and excites at 445nm.^43^

In this study, the sensitivity of GABase assay was further enhanced using resazurin (Sigma-Aldrich) which has a higher emission peak (587nm) when converted to fluorescent molecule resorufin. Four constructs with GAD binding-site mutations were used as knockdown positive controls: GAD65 [p.(Lys369Arg) & p.(Tyr425Phe)] and GAD67 [p.(Lys405Arg) & p.(Tyr434Phe)]. Less than 10% of GAD WT-activity was observed in all four knockdown controls which validates the high-sensitivity of the fluorescence-based resazurin-linked GAD enzymatic assay. GAD65 and GAD67 proteins (wild-type and mutant) were purified 24 hours after transfection and subjected to GAD enzymatic assay performed in 96-well plate format. 60µl of protein sample (up to 30µg of protein in lysates buffer containing 0.02mM pyridoxal 5′-phosphate (PLP), and 1mM 2-(2-Aminoethyl) isothiourea dihydrobromide (AET) in 100mM sodium phosphate (pH 7.2) was added to 60μl of reaction buffer (0.2mM PLP, 0.5mM DTT and 10mM glutamate) and incubated for up to 8 hours at 37°C. Reaction buffer without PLP was also used as control. Samples were centrifuged for 15min at 1000g to eliminate protein.

Aliquots of samples were then subjected to resazurin assay for quantifying GABA. The resazurin assay was performed using modified methods from previously described studies.^44^

GABA in each sample was measured using a freshly made resazurin solution containing 0.1 mM nicotinamide adenine dinucleotide (NAPD), 5 mM α-Ketoglutaric acid, 0.01 mM DTT, 0.05 U/ml GABase, 0.05 U/ml diaphorase, 6.25 µM resazurin in 100 mM Trisbase (pH 8.8). 10μl of samples from the GAD assay were added to a 96 well plate and then 90μl of the resazurin solution was added, prior to 1 hour incubation at RT protected from light. The fluorescent signal was measured using a FLUOstar Optima microplate reader (BMG Labtech) with the following setting: a 544 nm excitation/590 nm emission filter, gain 1327 and 10 flashes per well. GAD enzymatic activity assay standard curves are presented in **Supplementary Fig. 2**.

### Minigene splice-site assay

An *in vitro* splicing assay was used to analyse the splicing outcomes for *SLC6A1* IVS3+1G>A.^41^ The genomic region encompassing exons 1 to 4 of *SLC6A1* (4.155Kb) was PCR-amplified from P1 (**Table 1**) and control DNA using primer sets containing *Xhol* / *SacII* restriction sites: forward primer (5’-CACAGCTCGAGCTTCTTTCTTGCTCACC-3’), reverse primer (5’-CTATCCGCGGTCCAAC TCTTTTTGGGGAG-3’). PCR products were digested with *Xhol* and *SacII* and cloned into an exon-trapping (pET) vector (MoBiTec, Germany). WT and variant vectors were then transfected into HEK293 cells using Turbofectin^TM^ (OriGene). Following 24 hours, total RNAs were isolated using RNeasy (Qiagen, UK) based on the manufacturer’s protocol. 1µg of total-RNA was reverse-transcribed using SuperScript III kit (Invitrogen, UK) and PCR amplified using primer sets supplied with the Exontrap assay kit. PCR products were analysed on 2% (w/v) agarose gels and purified PCR products were sequenced.

**Table 1:** Clinical and phenotypic information in 31 GGE and Focal cases with GABAergic variants. Data was assembled from international tertiary epilepsy centers and all cases diagnosed by an experienced epileptologist and their teams. N/A: not applicable, GGE: genetic generalized epilepsy, TLE: temporal lobe epilepsy, CAE: childhood absence epilepsy, MAE: myoclonic astatic epilepsy, JME: juvenile myoclonic epilepsy, JAE: Juvenile absence epilepsy, FS: febrile seizures, FS+: febrile seizures plus, GTCS: generalized tonic-clonic seizures, EEG: electroencephalogram, MJ: myoclonic jerks, ASM: anti-seizure medication, ID: intellectual disability. A list of publications is presented in **Supplemental Table 4** when cases have featured in other studies or large consortia without any diagnostic or personalised outcomes.

| Case ID | P1 | P2 (digenic) | P3 | P4 | P5 |
| --- | --- | --- | --- | --- | --- |
| **Gene** | *SLC6A1* | *SLC6A1 / GAD2* | *SLC6A1* | *SLC6A1* | *SLC6A1* |
| **Variant** | IVS3 +1G>A | p.(Met179Val)<br>/ p.(Ser527Leu) | p.(Met179Val) | p.(Met555Val) | p.(Ser594Alafs*22) |
| **Age, Gender, Ethnicity** | *Removed* | *Removed* | *Removed* | *Removed* | *Removed* |
| **Electroclinical Diagnosis** | CAE | CAE | CAE | Possible CAE<br>(definite EEG changes) | CAE |
| **Family History** | *Removed* | *Removed* | *Removed* | *Removed* | *Removed* |
| **Seizure Types** | Absence | Absence, GTCS | Absence | Possible absences | Absence |
| **Seizure Onset (age in years)** | *Removed* | *Removed* | *Removed* | *Removed* | *Removed* |
| **Developmental Considerations after onset** | Difficulty Reading | Abnormal | ID | ID | ID |
| **EEG** | 3Hz Spike and Wave | 3Hz Spike and Wave and 3Hz Spike, polyspike and slow-waves | Spikes, polyspike and slow waves, 3-4Hz, photosensitive | Frequent paroxysms of sharp wave and spike and slow wave | Spikes and polyspike and wave |
| **ASM Medication History** | Unknown | Drug Refractory | Drug Refractory | VPA, unknown effectiveness | Drug Refractory |
| **Other Considerations.** | N/A | N/A | N/A | N/A | N/A |

| Case ID | P6 | P7 | P8 | P9 | P10 |
| --- | --- | --- | --- | --- | --- |
| **Gene** | *SLC6A11* | *SLC6A11* | *SLC6A11* | *SLC6A11* | *SLC6A11* |
| **Variant** | p.(Asn160Lys) | p.(Val256Leu) | p.(Asp295Asn) | p.(Arg436Gln) | p.(Arg597Trp) |
| **Age, Gender, Ethnicity** | *Removed* | *Removed* | *Removed* | *Removed* | *Removed* |
| **Electroclinical Diagnosis** | Right Lesional TLE | JAE | MAE | JAE | CAE to JME |
| **Family History** | *Removed* | *Removed* | *Removed* | *Removed* | *Removed* |
| **Seizure Types** | Temporal lobe seizures | Absence | febrile seizure plus, convulsions, myoclonic-atonic seizures, monthly absence, non-convulsive status | Absence | Absences (in childhood) GTCS; myoclonus as teenager; non-convulsive status |
| **Seizure Onset (age in years)** | *Removed* | *Removed* | *Removed* | *Removed* | *Removed* |
| **Developmental Considerations after onset** | None | None | mild-moderate ID | None | ID |
| **EEG** | Onset from Right TLE - Ictal EEG | 3.5-4.5 Hz Gen Spike and wave during hyperventilation, accompanied by a clinical absence. | slow background, generalised epileptiform activity; | Not available | 3hz Gen Spike & Wave, polyspike with slow wave discharges |
| **ASM Medication History** | Drug Responsive | Drug Responsive | Drug Refractory | Drug Responsive | Drug Refractory |
| **Other Considerations** | Underwent right temporal lobectomy | Born premature | Myasthenia Gravis (Thymectomy, 2000), Hypotonic, minor auricular deformity was noted (**Supp. Table 5**)<sup>1-3</sup> |  |  |

| Case ID | P11 | P12 | P13 | P14 | P15 |
| --- | --- | --- | --- | --- | --- |
| **Gene** | *GAD2* | *GAD2* | *GAD2* | *GAD2* | *GAD2* |
| **Variant** | p.(Cys45Phe) | p.(Pro153Gln) | p.(Pro153Gln) | p.(Pro153Gln) | p.(Pro153Gln) |
| **Age, Gender, Ethnicity** | *Removed* | *Removed* | *Removed* | *Removed* | *Removed* |
| **Electroclinical Diagnosis** | GGE | JAE | CAE | CAE/JAE | Early onset CAE, Occipital seizures |
| **Family History** | *Removed* | *Removed* | *Removed* | *Removed* | *Removed* |
| **Seizure Types** | Infrequent GTCS | Absence, absence status and GTCS | Absence and GTCS (rare) | Absence only | Febrile seizures, Focal occipital seizures, GTCS, visually induced Absences |
| **Seizure Onset (age in years)** | *Removed* | *Removed* | *Removed* | *Removed* | *Removed* |
| **Developmental Considerations After Onset** | None | None | Low average IQ | ID | Mild ID |
| **EEG** | 4.5-5Hz Gen spike & wave - polyspike & wave; photosensitive | Generalised spike & wave | 3Hz Generalised spike and Wave, sleep deprived EEG showing generalised spike and wave and polyspike and wave in sleep. | 3Hz Generalised Spike and Wave | 3Hz GSW<br>Some focal posterior slowing. |
| **ASM Medication History** | Drug Responsive | Drug Responsive | Drug Refractory | Drug Responsive | Drug Refractory |
| **Other Considerations** | Case published in (Supp. Table 4)<sup>4,5</sup> |  | Required 3 drugs at one stage. Now in remission | Case published in (Supp. Table 5)<sup>6,7</sup> | Clinically photosensitive hypertension & diabetes during pregnancy; Induced labour/emergency C-section due to abruption; Resuscitation needed at birth, slow to breathe |

| Case ID | P16 | P17 | P18 | P19 | P20 |
| --- | --- | --- | --- | --- | --- |
| **Gene** | *GAD2* | *GAD2* | *GAD2* | *GAD2* | *GAD2* |
| **Variant** | p.(Pro153Gln) | p.(Pro153Gln) | p.(Pro153Gln) | p.(Pro153Gln) | p.(Pro153Gln) |
| **Age, Gender, Ethnicity** | *Removed* | *Removed* | *Removed* | *Removed* | *Removed* |
| **Electroclinical Diagnosis** | Generalized epilepsy, possibly JME with unclear onset of MJ | FS+ | JME | JME | CAE |
| **Family History** | *Removed* | *Removed* | *Removed* | *Removed* | *Removed* |
| **Seizure Types** | Myoclonic seizures, GTCS, FS | GTCS | GTCS | Myoclonic seizures, GTCS | Absence (from 4y), GTCS (14-27) |
| **Seizure Onset (age in years)** | *Removed* | *Removed* | *Removed* | *Removed* | *Removed* |
| **Developmental Considerations after onset** | None | Severe speech delay, dyspraxia and bilateral mild sensorineural hearing loss. | Speech delay | None | None |
| **EEG** | 4-6Hz polyspike & wave, photosensitive | R Centro-temporal spikes | Generalised spike & wave, polyspike & wave, at rest, HV & photic stimulation. Photosensitive | 5Hz Generalised spike & wave, polyspike & wave | Normal at rest, clinical & electrical "petit mal" evoked by HV (burst of spike wave activity) |
| **ASM Medication History** | Drug Responsive | Drug Responsive | Drug Responsive | Drug Refractory | Drug Refractory |
| **Other Considerations** | Cases previously in published study. See (Supp. Table 5)<sup>8,9</sup> | SCN1A in the family Cases previously in published study. See (Supp. Table 5)<sup>10</sup> | Submitted to the Epi4K Consortium and published in (Supp. Table 5)<sup>5</sup> |  | Mother miscarried twin early on |

| Case ID | P21 | P22 | P23 | P24 | P25 |
| --- | --- | --- | --- | --- | --- |
| **Gene** | GAD2 | GAD2 | GAD2 | GAD2 | GAD2 |
| **Variant** | p.(Pro153Gln) | p.(Pro153Gln) | p.(Pro153Gln) | p.(Pro153Gln) | p.(Pro153Gln) |
| **Age, Gender, Ethnicity** | *Removed* | *Removed* | *Removed* | *Removed* | *Removed* |
| **Electroclinical Diagnosis** | CAE | Generalized epilepsy with intellectual disability | CAE | CAE | CAE |
| **Family History** | *Removed* | *Removed* | *Removed* | *Removed* | *Removed* |
| **Seizure Types** | Absences | Absences, GTCS | Absence | Absence | Absence |
| **Seizure Onset (age in years)** | *Removed* | *Removed* | *Removed* | *Removed* | *Removed* |
| **Developmental Considerations after onset** | None | Moderate-Severe ID: Overall cognitive ability currently in extremely low range | None | None | None |
| **EEG** | Generalised spike & wave | Generalised polyspike & wave. Photosensitive | 3-3.5Hz spike & wave | Prolonged runs of 2.5Hz spike & wave, photosensitive | 3Hz spike, polyspike & waves, photosensitive |
| **ASM Medication History** | Drug Refractory | Drug Refractory | No medication | Drug Responsive | Drug Responsive |
| **Other Considerations** |  | 2p16.3 deletion |  |  |  |

| Case ID | P26 | P27 | P28 | P29 | P30 | P31 |
| --- | --- | --- | --- | --- | --- | --- |
| **Gene** | *GAD2* | *GAD2* | *GAD2* | *GAD1* | *GAD1* | *GAD1* |
| **Variant** | p.(Pro153Gln) | p.(Pro153Gln) | p.(Ile575Thr) | p.(Arg89Gly) | p.(Tyr127Phe) | p.(Tyr127Phe) |
| **Age, Gender, Ethnicity** | *Removed* | *Removed* | *Removed* | *Removed* | *Removed* | *Removed* |
| **Electroclinical Diagnosis** | CAE | Lesional TLE | CAE/JAE | CAE | JAE to JME | FS |
| **Family History** | *Removed* | *Removed* | *Removed* | *Removed* | *Removed* | *Removed* |
| **Seizure Types** | Absence, GTCS | Temporal lobe seizures | Absence, GTCS | Absence | Absence, myoclonic seizures, GTCS | FS |
| **Seizure Onset (age in years)** | *Removed* | *Removed* | *Removed* | *Removed* | *Removed* | *Removed* |
| **Developmental Considerations after onset** | None | None | None | None | None | None |
| **EEG** | 3Hz spike & wave | NA | Double or multiple spikes with polyspike & wave | 3-4Hz spike & wave, photosensitive | Generalised spike & wave, photosensitive | Normal |
| **ASM Medication History** | Drug Responsive | Drug Responsive | Drug Refractory | Drug Responsive | Drug Refractory | Not on ASM's |
| **Other Considerations** |  | Resection of pilocytic astrocytoma in 1990's |  |  | Submitted to the Epi4K Consortium (Supp. Table 5)<sup>5</sup> |  |

### Statistical analysis

Datasets were analyzed using GraphPad Prism (GraphPad Software Inc) and expressed as mean ± standard error of mean (SEM). Statistical significance was determined by Student’s *t*-test and considered to be significant at p<0.05.

## Results

### Novel and ultra-rare Variants in *SLC6A1, SCL6A11, GAD1* and *GAD2*

Following genetic analysis of 4 GABAergic genes *(SLC6A1, SCL6A11, GAD1* and *GAD2)* in 708 people with epilepsy, 19 novel or rare variants (MAF <0.01) were taken forward to functional studies. Based on the functional analysis, 15 out of 19 variants identified in 31 cases were regarded as pathogenic representing 4.4% of the cohort (**Table 1, Supplemental Table 2, 3**). The remaining four variants did not reach levels of functional significance, closely resembling WT activity levels (**Supplemental Table 4**). The 15 pathogenic variants also showed potentially damaging outcomes based on phylogenetic alignment, mutation-prediction software, and molecular modelling (**Supplemental Table 2, Supplementary Figure 3, 4, 5 and 6**). One case (P2) has very rare digenic *SLC6A1* and *GAD2* variants. Identification of *SLC6A11* (GAT3) and *GAD2* (GAD65) are novel findings in epilepsy, whilst *SLC6A1* (GAT1) and *GAD1* (GAD67) variants add to the compendium of published epilepsy genes with new mechanistic functional outcomes.^25–32^

### Loss of Function and Reduced Transporter Activity for GAT1 and GAT3 Transporters

Nine heterozygous *SLC6A1* and *SLC6A11* variants were discovered in 10 unrelated people (**Table 1, Supplemental Table 2, Figure 1**). Four very rare *SLC6A1* variants were discovered in five people with epilepsy (four of whom had GGE) including one essential splice-site, one **nonsense** frameshift and two missense variants. *SLC6A11* missense variants were discovered in five unrelated GGE cases, linking the GAT3 transporter to GGE as a novel finding.

**Figure 1:**
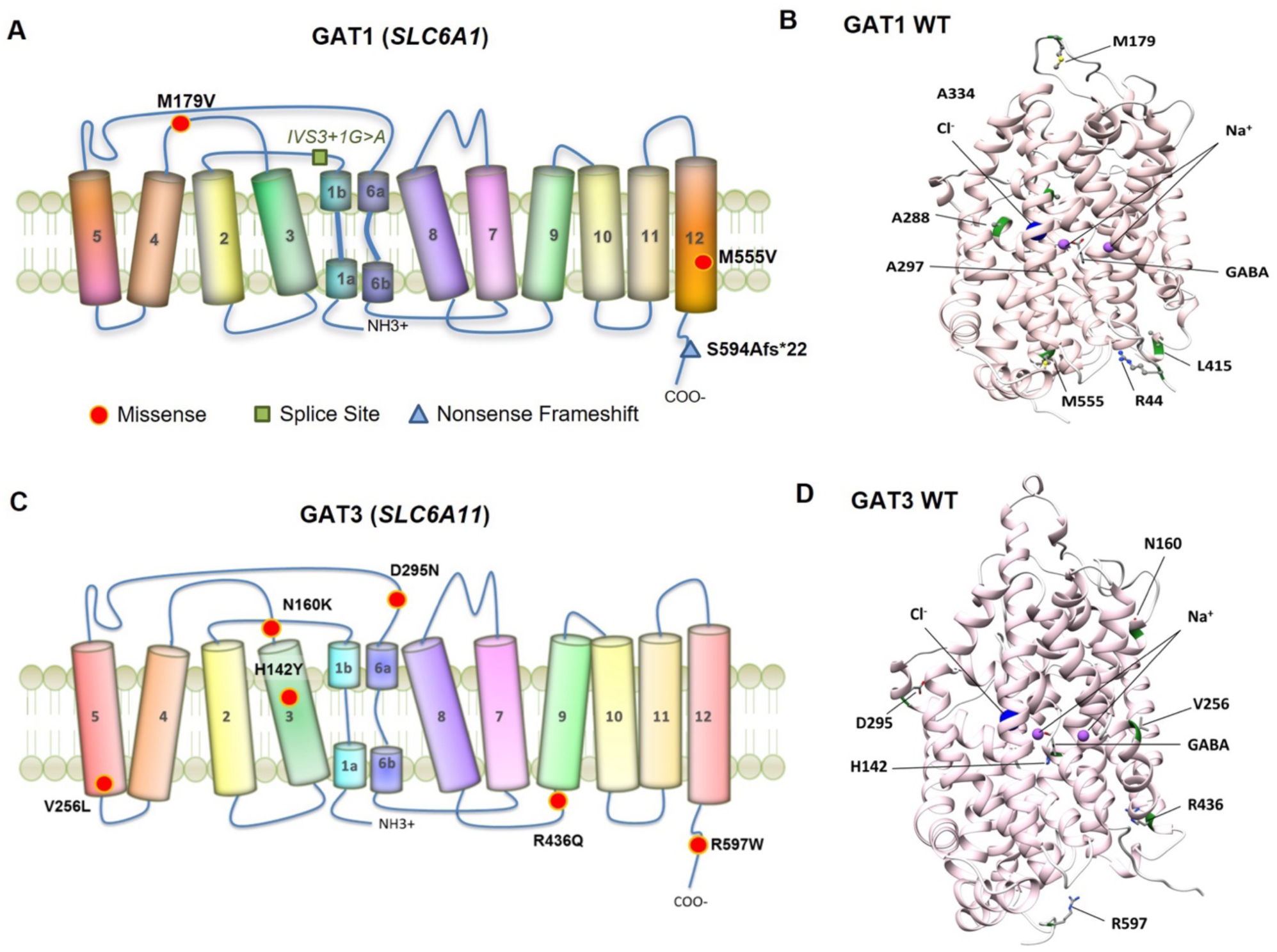
Novel and ultra-rare gene-variants found in GABA Transporter genes *SLC6A1* (GAT1) and *SLC6A11* (GAT3). **(A and C)** show the 2-D representation of GAT1 and GAT3 topology and organisation of the transmembrane domains. Overlaid are the approximate position of each gene-variant detected in this study with missense changes (red spots), splice site changes (green squares), and frameshifts mutations (blue triangle). **(B and D)** Molecular modelling of GAT1 and GAT3 wild-type structures, showing the 3D location of mutated amino acids, and the binding of GABA, Cl^-^ and Na^+^ ions.

Nine S*LC6A1* / *SLC6A11* variants from this study plus an additional seven *SLC6A1* MAE variants from Carvill *et al* (2015)^25^ were analysed through isotopically labelled GCMS GABA transport activity method (**Figure 2**). All seven MAE-linked *SLC6A1* variants displayed complete loss-of-function outcomes in comparison to WT GABA transport activity (**Figure 2A**). The nonsense / frameshift variants (*SLC6A1*) c.452delT p.(Leu151ArgfsTer35), c.578G>A p.(Trp193Ter), and c.1369_1370delGG p.(Gly457HisfsTer10) had loss of protein expression and created a range of premature truncated products (**Figure 2B**).

**Figure 2:**
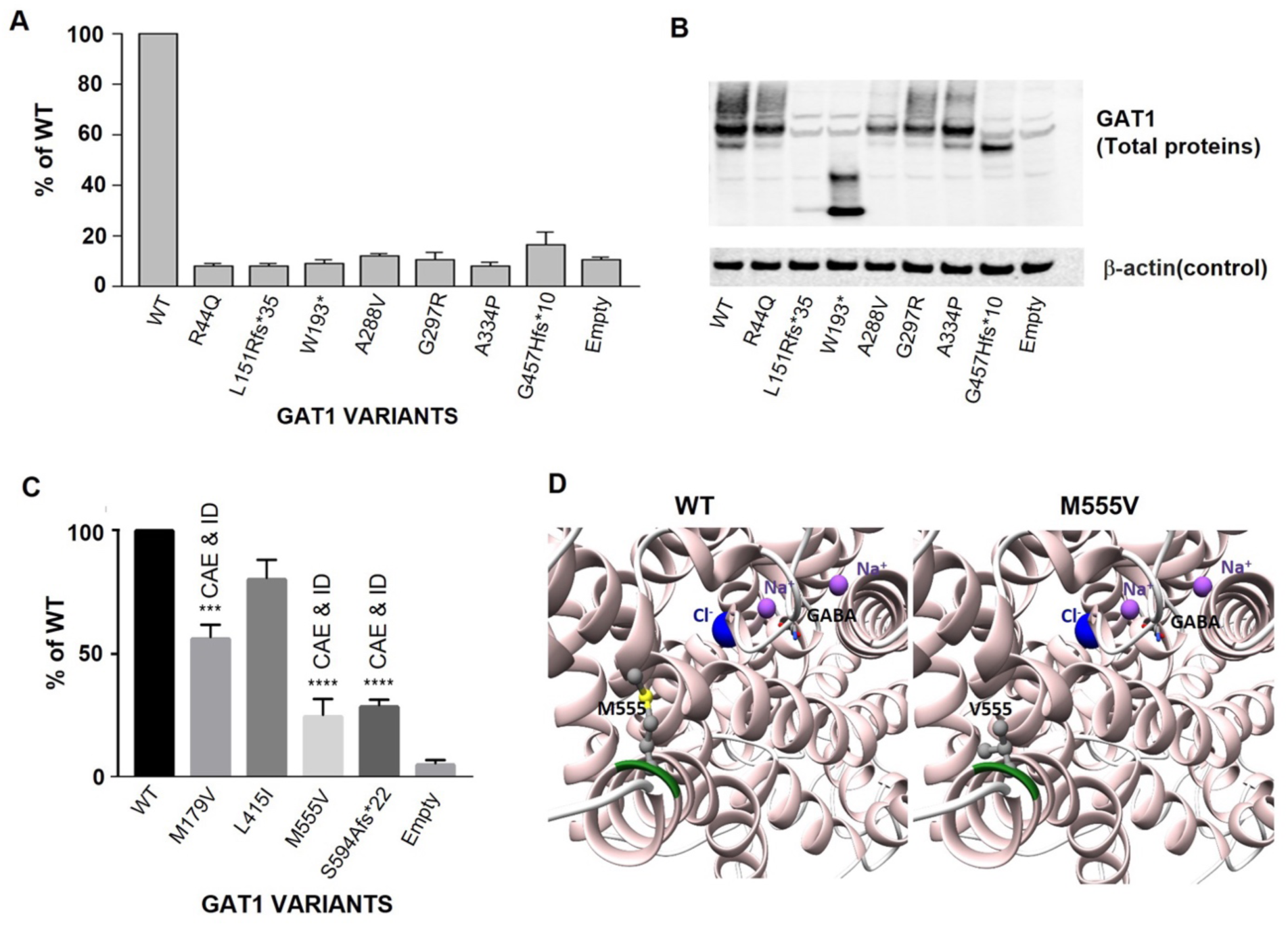
Functional Analysis of *SLC6A1* (GAT1) Variants. **(A)** LoF myoclonic astatic epilepsy (MAE) mutations^24^ tested through the GABA transporter assay and resulting in total activity knockdown in comparison to WT. **(B)** A total protein extraction Western blot of the Carvill *et al (2015)* mutations with nonsense and frameshift mutations (lanes 3, 4 and 8) resulting in truncated proteins. **(C)** GABA transporter assay on *SLC6A1* variants found in this study with observations of 45%–75% reduction in GAT1 mediated GABA transport activity relative to GAT1 WT and empty vector. The GGE syndrome is labelled above each column-bar corresponding to gene-variants that reached significance as indicated (p-value of <0.001 as *** and p value of <0.0001 for ****). **(D)** Molecular modelling of GAT1 protein comparing WT structure with GAT1 c.1663 A>G p.(Met555Val) structure. This shows the changes to the cytoplasmic exit of the pore region, and potential disruption to the transport of ions and GABA binding sites.

From this study, two (*SLC6A1*) variants c.1780delA p.(Ser594Alafs*22), and c.1663 A>G p.(Met555Val) also showed significant reduction of GAT1 activity (**Figure 2C**), whilst a partial, but significant, reduction of transporter activity was detected for the p.(Met179Val) missense variant. 3D modelling of GAT1 p.(Met555Val), located at the cytoplasmic exit of TM12, predicts effects on the pore region and transport of ions (**Figure 2D)**. We also conducted *in vitro* splicing analysis on the novel (*SLC6A1)* c.471+1G>A variant, confirming the skipping of exon 3 from RT-PCR transcripts creating a frameshift and premature truncated protein (**Figure 3A-C**). All five GGE cases had co-morbid ID or developmental delay, consistent with previously published studies (**Table 1**).^25–30^

**Figure 3:**
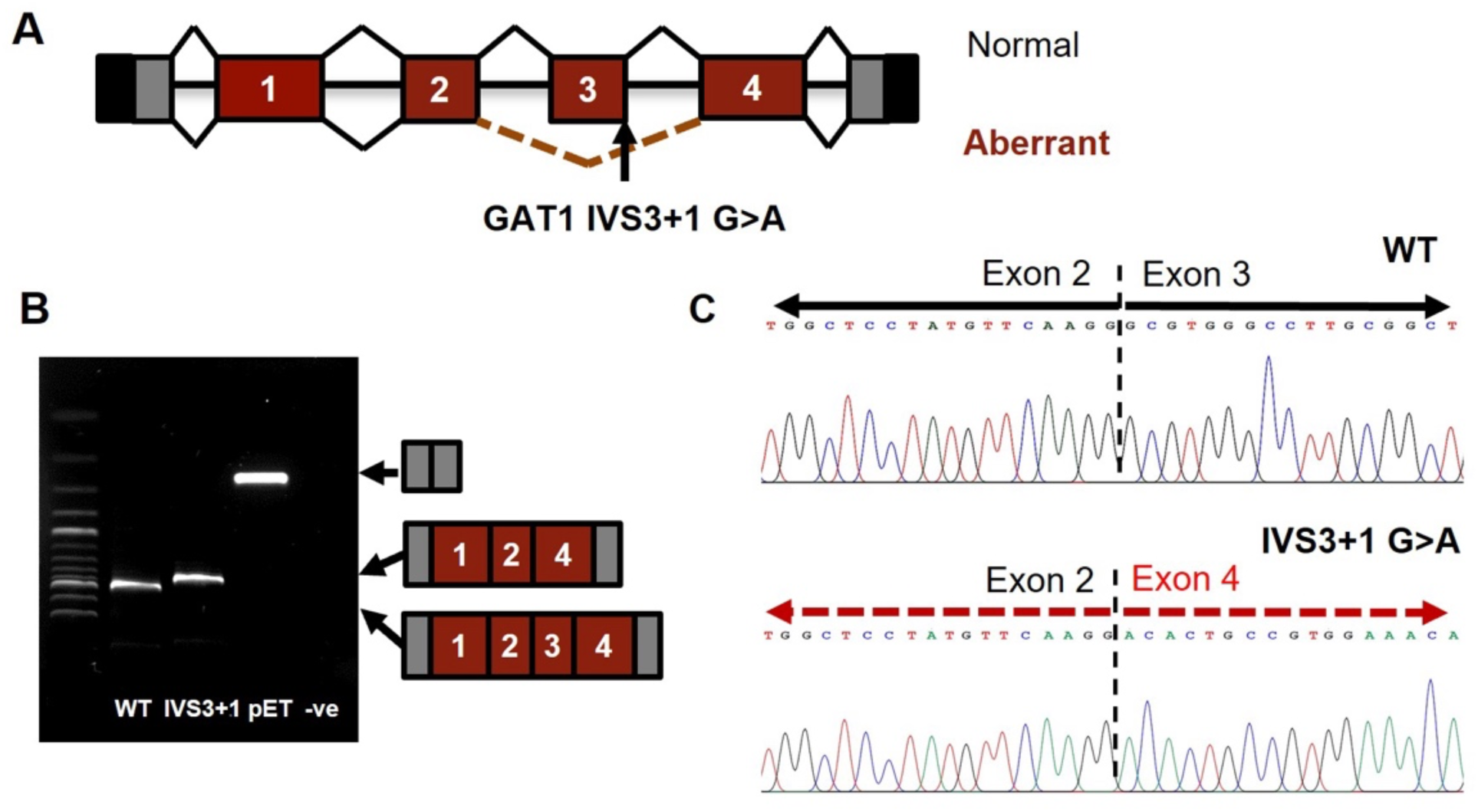
Splice site assay of *SLC6A1* IVS3+1G>A. **(A)** Splice-site analysis of *SLC6A1* c.471+1G>A, IVS3+1 G>A (arrow) reveals a predicted alternate splicing pattern. **(B)** RT-PCR from RNA isolated from the *in vitro* splice-site assay showing a PCR-size difference between WT and IVS3+1G>A. **(C)** Sanger sequencing of the *SLC6A1* c.471+1G>A, IVS3+1G>A RT-PCR reveals a precise transcript deletion of exon 3 with WT RT-PCR resulting in normal splicing patterns. The end of *SLC6A1* exon 2 joins the start of *SLC6A1* exon 4, which excludes exon 3, and creating a frameshift protein truncation event.

Five *SLC6A11* missense variants displayed significantly reduced GABA transport activity compared to WT **(Figure 4A)**. (*SLCA11*) c.883G>A p.(Asp295Asn) had the most reduced transport activity and is associated with a severe MAE phenotype reminiscent of GAT1-mediated loss-of-function MAE. Variants (*SLCA11*) c.480C>G p.(Asn160Lys), c.766G>T p.(Val256Leu), c.1307G>A p.(Arg436Gln), and c.1791C>T p.(Arg597Trp) showed a 45–50% reduction in GABA transporter activity across a range of GGE syndromes plus one focal case with temporal lobe epilepsy (TLE) **(Table 1)**. Cell-surface biotinylation experiments revealed that the degree of reduced GAT3 cell-surface trafficking correlates with the severity of reduced GABA activity and the consequential GGE phenotype (**Figure 4B**). Molecular modelling indicates p.(Asp295Asn), located critically at the extracellular entry of TM6, severely perturbs the binding and passage of GABA through the pore by loss of alignment of side chain groups that direct the ligand to the core of the protein (**Figure 4C**).

**Figure 4:**
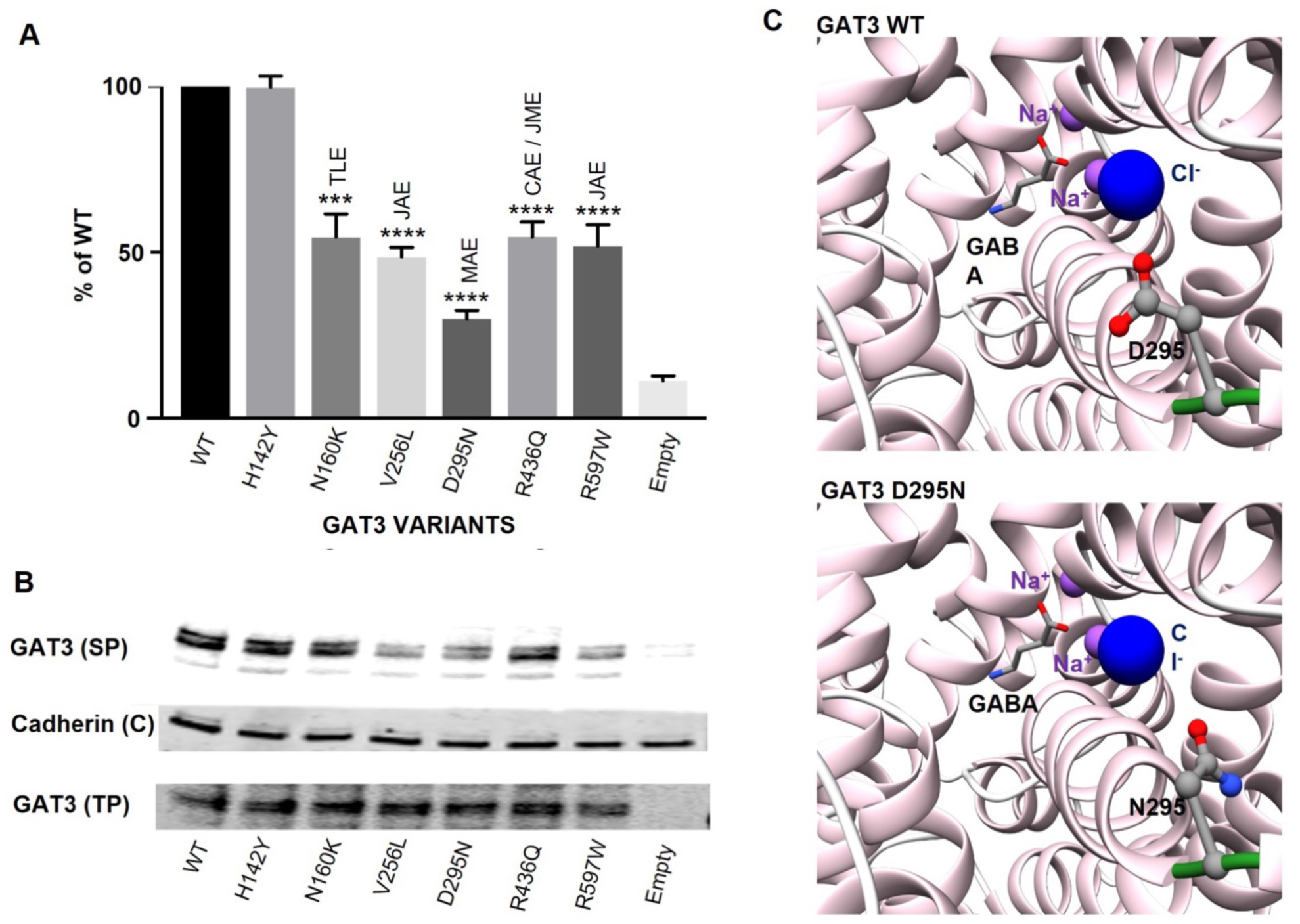
Functional Analysis of *SLC6A11* (GAT3) Variants. **(A)** The GABA transporter assay on *SLC6A11* variants revealed a 40%–70% reduction in GAT3 mediated GABA transport activity relative to GAT3 WT and empty vector. GGE syndromes and levels of significance are indicated above the column-bars (p-value of <0.001 as *** and p value of <0.0001 for ****). **(B)** Cell-surface biotinylation of the GAT3 variants shows reduced cell-surface proteins for the three severely impacting variants when compared to whole-cell levels and control proteins (cad). SP: surface protein, C: control, TP: total protein. **(C)** A molecular modelling example of (GAT3) c.883G>A p.(Asp295Asn) showing the disruption of side chain orientation at the extracellular entry to the passage of ions and GABA in comparison to GAT3 WT.

### Reduced GAD65 Activity and Gain-of-Function outcomes for GAD67

Six highly conserved, rare *GAD1* and *GAD2* variants were discovered in 22 unrelated people with epilepsy (19 with GGE) (**Table 1**, **Figure 5A, Supplemental Table 2, 3**). Four N-terminal and C-terminal (*GAD2)* missense variants were found in 19 GGE cases, with two variants c.134G>T p.(Cys45Phe), c.1725T>C p.(Ile575Thr) being private changes, c.1580C>T p.(Ser527Leu) is digenic with (*SLC6A1*) c.535A>G p.Met179Val (p2), and (*GAD2*) c.358C>A p.Pro153Gln was found in 16 cases. We included p.(Pro153Gln) in functional studies because the bioinformatics predicted a damaging outcome, and the frequency in epilepsy cases (2%) was much higher than the ethnically matched control populations (0.84%). The molecular modelling of p.(Pro153Gln) indicates significant changes in orientations of side chains along a flexible loop region containing Gln190 and Ser192, leading to the loss of ligand hydrogen bonding to Ser192 (**Figure 5B-D**).

**Figure 5:**
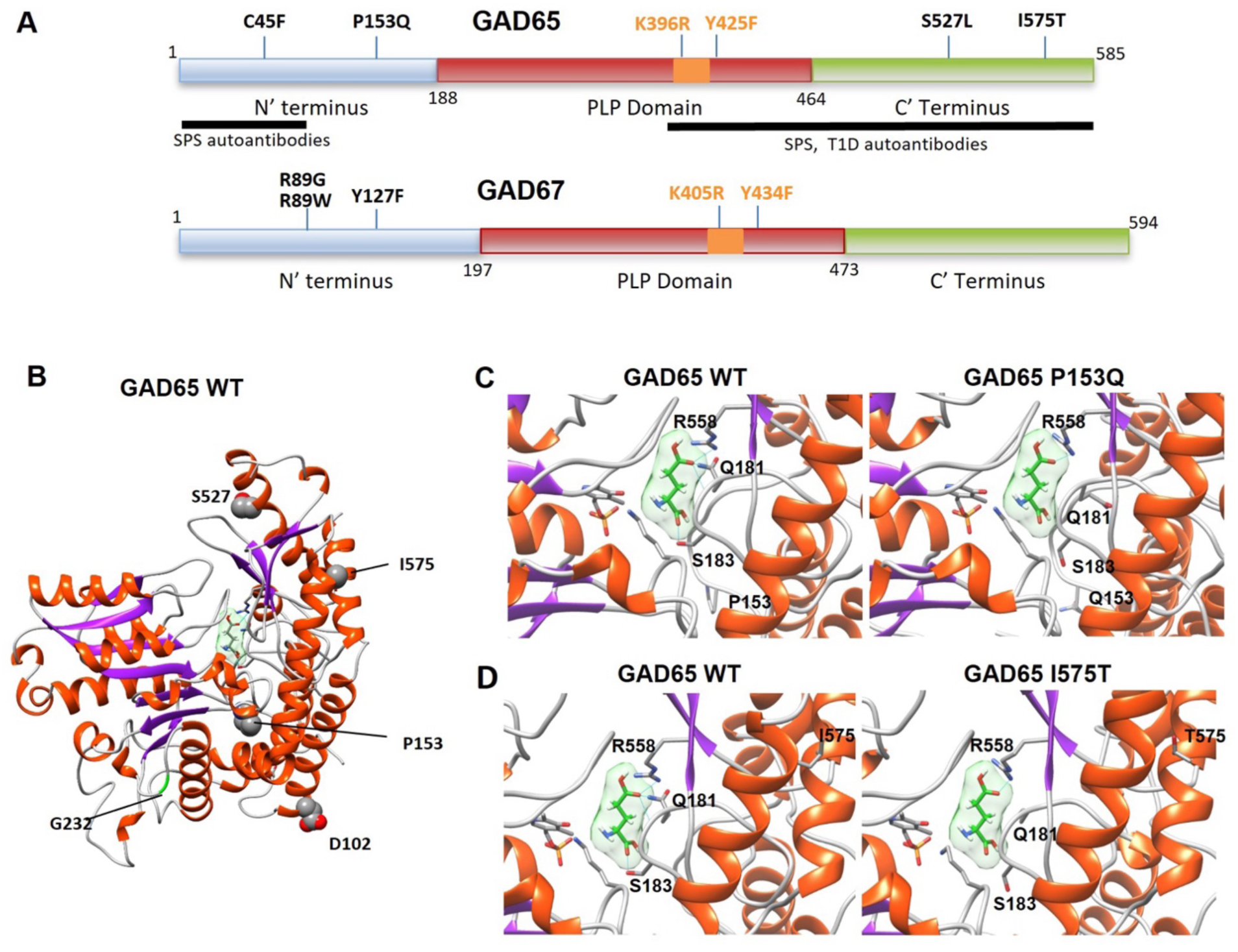
Novel and ultra-rare gene variants in *GAD1* (GAD67) & *GAD2* (GAD65). **(A)** an established GAD65 / GAD67 protein domain map with the N-terminus in blue, PLP domain in red, the PLP binding region in orange, and the C terminus in green. The position of the *GAD1* / *GAD2* variants within the GAD protein domains are displayed in addition to the location of the control *GAD* knockdown control mutations (p.K396R, p.Y425F, p.K405R and p.Y434F) represented in orange font. **(B-D)** Molecular modelling of GAD65 WT and gene variants displaying the effects of variants on the local structure of the ligand binding pocket and effects on interactions.

In addition, two ultra-rare *GAD1* variants were discovered in three people with epilepsy. The (*GAD1*) c.265C>G p.(Tyr127Phe) variant showed phenotypic discordance, identified in one person with a familial GGE syndrome, whilst the other had febrile seizures in a Genetic Epilepsy with Febrile Seizures plus (GEFS+) family. Recessive or bi-allelic *GAD1* cases were not identified in this cohort in contrast to recent reports in more severe developmental and epileptic enchepalopathy.^31–32^ Molecular modelling was not possible for *GAD1* variants because the GAD67 N-terminal region remains elusive to accurate structural predictions.

We used a florescence-based, resazurin-linked GAD enzymatic activity assay (**Figure S2**) to functionally assess the GAD variants. All four (*GAD2)* variants c.134G>T p.(Cys45Phe), c.358C>A p.(Pro153Gln), c.1580C>T p.(Ser527Leu) and c.1725T>C p.(Ile575Thr) displayed a significant loss-of-function reduction in GAD65 enzymatic activity compared to WT (**Figure 6A**). Loss-of-activity were observed in both GAD65 knockdown control variants. Analysis of GAD67 enzymatic activity showed that (*GAD1)* c.265C>G p.(Arg89Gly) and c.265C>G p.(Tyr127Phe) produced an excess of activity compared to WT, indicating a potential gain-of-function outcome (**Figure 6B**). Identified in this study, a c.265C>G p.(Arg89Trp) variant (**Supplemental Table 4**) displayed full WT activity whereas the GAD67 knockdown controls have a complete loss of GAD activity.

**Figure 6:**
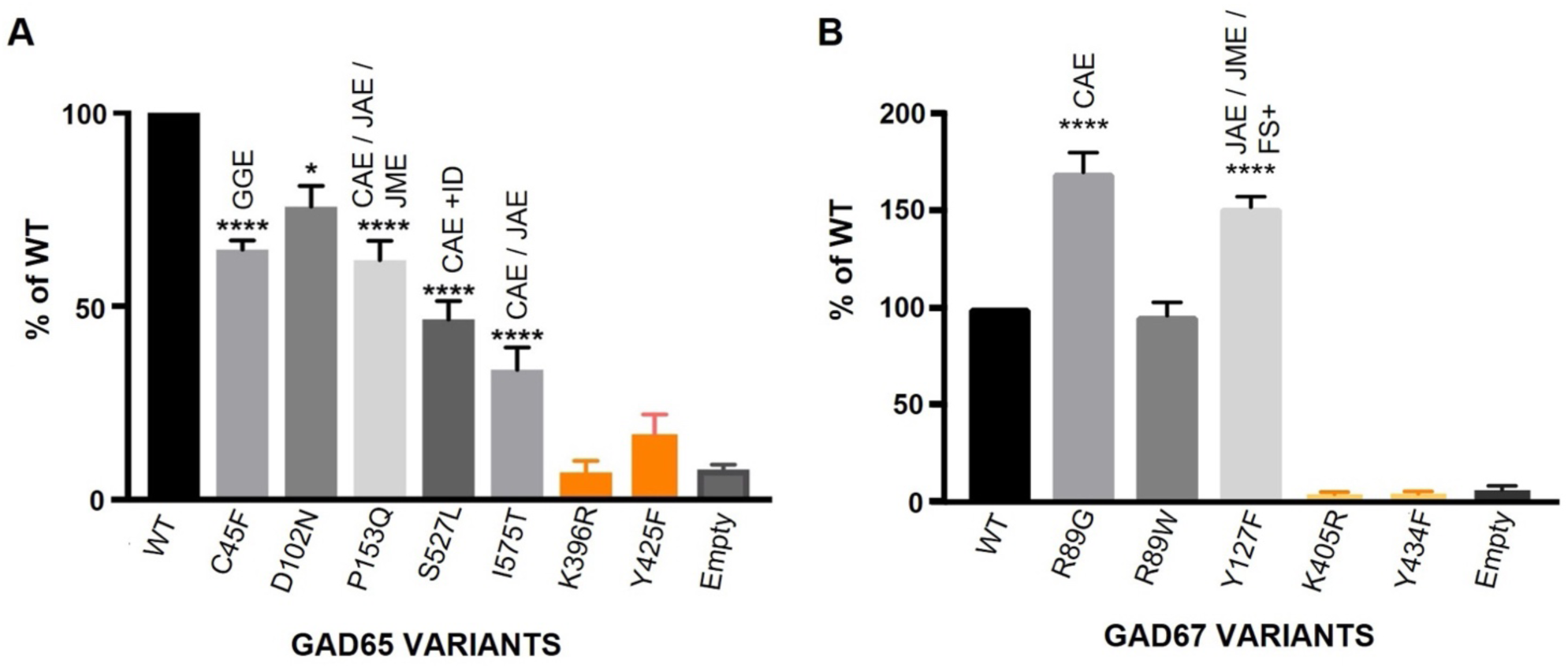
A novel and enhanced GAD enzymatic assay to functionally characterize the effect of GAD65 / GAD67 protein variation. **(A)** a GAD enzyme activity assay was developed to test *GAD2* variants versus WT and knockdown controls (K396R; Y425F; in orange). Results were confirmed in triplicate thereby minimizing interval limits and increasing confidence in significance levels. Corresponding GGE syndrome(s) were labelled above each variant activity columns (Table 1). Compared to wild-type activity levels, all GAD65 variants displayed highly-significant reduction in GAD65 enzyme activity. **(B)** the GAD enzyme activity assay also characterized *GAD1* variants versus WT and knockdown controls (K405R; Y434F; in orange). Results were confirmed in triplicate and the GGE syndrome(s) are labelled above the variant activity columns. Compared to wild-type activity levels, the GAD67 variants displayed highly-significant increase in GAD67 enzyme activity reflecting a possible gain-of-function effect.

#### Genotype / Phenotype Correlations Show Differences in Clinical Outcomes

The five individuals with *SLC6A1* mutations all had CAE, three of whom were drug refractory (**Table 1**). None had normal education attainment, which is atypical for CAE with challenges ranging from a specific reading difficulty to ID. The five with *SLC6A11* variants were discordant for epilepsy syndrome, one had a focal onset epilepsy (p6), one had MAE (p8) whilst the others had more typical GGE absence-predominant epilepsies; again, three were drug refractory (p6, p8, p10). 19 cases had *GAD2* variants, with 17 having GGE syndromes, one case with TLE and one unclassified GGE. Of the 17 GGE cases, two were confirmed JME, eight were drug-refractory and seven had educational needs ranging from mild to moderate ID. No educational challenges were reported in the three cases with *GAD1* variants.

Due to case ascertainment biases, we had an excess of cases with a notable family history - 21 in total (68%). Across the cohort with variants (n=31), two had TLE both of whom underwent lesional resections, two were part of GEFS+ families and the remainder had GGE syndromes: 15 with CAE, three with CAE/Juvenile Absence Epilepsy (JAE) indistinguishable, three with JAE, three with GGE without a clear syndrome, one JAE to JME, one CAE to JME and one with MAE. 50% were responsive to anti-seizure medications (ASMs) (n=17), whilst 42% were drug-refractory (n=15) with two cases having no drug-treatment recorded. Cases with either GAT1 or GAT3 variants with severe reduction in transporter activity have a more severe GGE as represented by seizure control and comorbidities, i.e. MAE compared to JAE. GGE cases with the most severe reduction GAD65-enzymatic activity are drug-refractory. Notably, the one case with a pharmacoresistant early-onset CAE (P2) has digenic ultra-rare *SLC6A1* / *GAD2* variants. The two gain-of-function GAD67 variants varied in presentation severity from a drug-refractory JME to drug-responsive CAE and a history of early-onset febrile seizures. Generally, individuals with GAD65 and GAD67 protein variants have no educational concerns.

## Discussion

This study has found 15 functionally validated heterozygous *SLC6A1, SLC6A11, GAD1* and *GAD2* variants in 31 people with epilepsy predominantly with GGE syndromes, including one digenic case (p2) and two focal TLE cases (p6, p27). Structural and functional analysis for GAT1, GAT3, GAD65 and GAD67 has revealed a profound detrimental effect on GABA transporter activity and biosynthetic enzymatic GABA production. Linking epilepsy with deleterious *SLC6A11* / *GAD2* variants plus new gain-of-function *GAD1* variants are novel findings, whilst new genotypes in *SLC6A1* and *GAD1* add to the growing consensus that these are major pathogenic GGE and developmental and epileptic encephalopathy (DEE) genes. We suggest that the new genes described in this study may not have a signal in larger epilepsy consortium outcomes because of more stringent MAF thresholds in NGS analyses (e.g. <0.001). To date, we have not detected any homozygous or bi-allelic cases reflecting a consistent bias towards heterozygous variants in the GABAergic genes as a common inheritance feature. With the recent exception of *GAD1* homozygosity, it is likely that homozygous GABAergic variants cause *in utero* lethality or debilitating life-limiting infantile encephalopathy.^45^

*SLC6A1* is now a recurrent diagnostic GGE gene where knockdown loss-of-function variants are a cause of DEE, characteristically an MAE whereas partial perturbation of GABA uptake activity is most likely causing less severe GGE syndromes that are drug-resistant and co-present with ID. This is supported by data from the Epi25 consortium showing *SLC6A1* has a strong gene-burden GWAS signal in GGE and DEE.^4,46^ Convergent evidence in human and animal models show that GAT1 loss-of-function results in increased tonic inhibition through excess extracellular GABA levels that remain in the synaptic cleft.^17–19^ This may cause absence seizures in children through cortical inactivation of the sensitive and rhythmic cortico-thalamo-cortical circuitry.^47–48^ Historical GAT1 knockout mice display tremors, ataxia and anxiety whist more recent haploinsufficient knock-in murine models display epilepsy with increased 5–7Hz spike-wave discharges and absence seizures.^49–52^ Scale, complexity, brain-region sensitivities / specialisations, and a host of other anthro-biological reasons may limit translational relevance. For example, it is not explained why GAT1 reduction or loss-of-function is biologically causing ID and cognitive effects in most heterozygous carriers. This may be a direct effect of *in utero* brain development, infantile / perinatal connectome pruning, developmental GABA interneuron influences, seizure duration / frequency or status epilepticus.^53–54^

Functionally validated *SLC6A11* mutations have not previously been linked to GGE where reduction of GAT3 mediated GABA uptake activity is linked to a wide spectrum of drug-resistant GGE syndromes and 1 TLE post-surgical remission case. In contrast with *SLC6A1* carrier presentation, there is no evidence of consistent pattern of cognitive co-morbidity which may underpin some of the GAT3 biological uniqueness in brain development or cortico-thalamic physiology. GAT3 demonstrates the same trending principle where a sliding scale of transporter activity reduction is linked to more severe GGE syndrome presentation. GAT3 p.Asp295Asn is a primary example where severe reduction in transporter activity manifests as MAE and the only GAT3 variant causing mild-moderate ID. GAT3-mediated partially reduced GABA activity is causing a spectrum of relatively milder GGE syndromes. GAT3 transporters generally localize to astroglial cells, however, in the thalamus they are found on cortical-thalamic inhibitory neurons indicating a specialized role in an epileptogenic-sensitive junctional complex of the subcortical human forebrain.^15–17^ GAT3 dysfunction has been linked to other disorders, for example expression studies of GAT3 show downregulation in IPSC astrocytes in Alzheimer Disease, murine neuromodulation of striatal astrocytes in obsessive-compulsive-like behaviour and tonic inhibition in external globus pallidus neurons in Parkinsonian rodents.^13, 55–58^

For GAD isoform enzymes, an enhanced-sensitivity GABA enzymatic assay has revealed a variable loss-of-function in activity for GAD65 and a gain-in-function activity for GAD67 in a spectrum of GGE presentations. This, for the first time, has directly linked GAD65 to human epilepsy syndromes after some considerable biological evidence underpinning their pathogenic candidacy.^21–23^ ^59–61^ *Gad65* -/- mice exhibit increased anxiety and fear responses, and have a significantly lowered seizure threshold with consequences for GABA production / glutamate storage. Auto-antibodies targeting GAD65 are associated with stiff-man syndrome, insulin-dependent diabetes mellitus and several neurological phenotypes including epilepsy. GAD65 is the reactive biosynthesis isoform when more GABA is required and is rapidly activated and deactivated upon PLP binding. Therefore, the lower GAD65 enzymatic activity observed in this study may cause a quantal GABA deficit in synaptic vesicle filling during presynaptic release.

Unlike the GAT1 and GAT3 observations, there is no trend established between GAD65 knock-down levels and epilepsy syndrome severity. However, it remains a strong link to clinically important electroclinical syndromes. This study shows that GAD65 p.(Pro153Glu) may be directly pathogenic or given its high frequency in the population, could be regarded as a susceptibility variant. The digenic GAT1/GAD65 case observed in this study (P2) is also interesting given the double hit of pathogenic presynaptic / astroglia pathology with a CAE with ID / drug-resistance phenotype.

GAD67, in contrast, is responsible for steady-stream constitutive production of neuronal / vesicular GABA and is essential for normal neuronal function and development.^21^ Recently, bi- allelic loss-of-function *GAD1* variants have been discovered in a developmental epileptic encephalopathy and often results in early childhood death.^31–32^ In our study, we have found two heterozygous *GAD1* variants in three epilepsy cases that exhibit a novel GAD67 enzymatic overactivity and a possible gain-of-function effect. This may indicate that *GAD1* heterozygosity could be causing childhood epilepsy syndromes but biallelic loss of function are creating DEE. However, how over-active GAD67 enzymatic activity is epileptogenic is theoretically the production of excess quantal GABA creating instabilities in vesicular filling / presynaptic release or could upset the delicate GAD65/GAD67 quantal ratios or switch-on/switch-off activity. Further *GAD1* variant discoveries will help validate this gain-of-function expression pattern. Other gain-of-function neuro-enzymopathies include *ADCY5*-related dyskinesia, *SOD1* in ALS, *GLUD2* in Parkinson disease and *ODC1* in neurodevelopmental disorders.^62–66^

In this study, we have demonstrated that *SLC6A11* and *GAD2* are new causative epilepsy genes, with further evidence confirming *SLC6A1* and *GAD1* as established diagnostic genes. Furthermore, how this study translates into *in vivo* causality within the GABAergic neurotransmission will occupy future workstreams. Thalamocortical networks have a distinct reliance on GABA networks / physiology and could be especially susceptible to quantal GABA fluctuations. This may lead to oscillatory cortico-thalamic disturbance manifesting as prenatal connectome disturbance, childhood absence / atonic seizures and core components of the GGE syndrome spectrum.^67–71^

We have conclusively added to the presynaptic GABAergic drivers of epilepsy syndromes and further confirm that GABA network perturbation is an important factor for epileptogenesis.

## Data Availability

All data produced in the present work are contained in the manuscript

## Acknowledgments

We would like to thank all the patients and their families for participating in research and providing consented DNA samples.

## Author Contributions

SK.C, R.J.H and M.I.R, were responsible for the conceptualization of the research. SK.C defined the direction of the research, designing and performing the functional platforms with assistance from E.D for mass spectrometry assays. SK.C supervised the genetics team (C.A.H, A.V.D, A.H, J.S) and funded the research through a ERUK Fellowship. J.G.L.M performed 3D bioinformatic modelling analysis. Clinical interpretation was provided by R.H.T, S.T.B and I.E.S; whilst S.F.B, I.E.S, S.T.B, P.S.B, and K.V.E provided the DNA and clinical details for the GGE cases from Australia, New Zealand and London. SK.C, R.H.T, J.G.L.M and M.I.R wrote the manuscript with all authors contributing to the final version.

## Data Availability

New genetic findings will be submitted to NIH dbSNP, and gnomAD database (see methods).

## Supplementary Material

Supplementary material is available online with 6 supporting Figures and 5 Tables.

