## Supplementary Information for "Pathogenic ultra-rare variants in *SLC6A1*, *SLC6A11*, *GAD1* and *GAD2* are new & recurrent GABAergic loci for GGE syndromes"

#### Supplementary Figures 1-6

#### Supplementary Tables 1-5

### Supplementary Figures

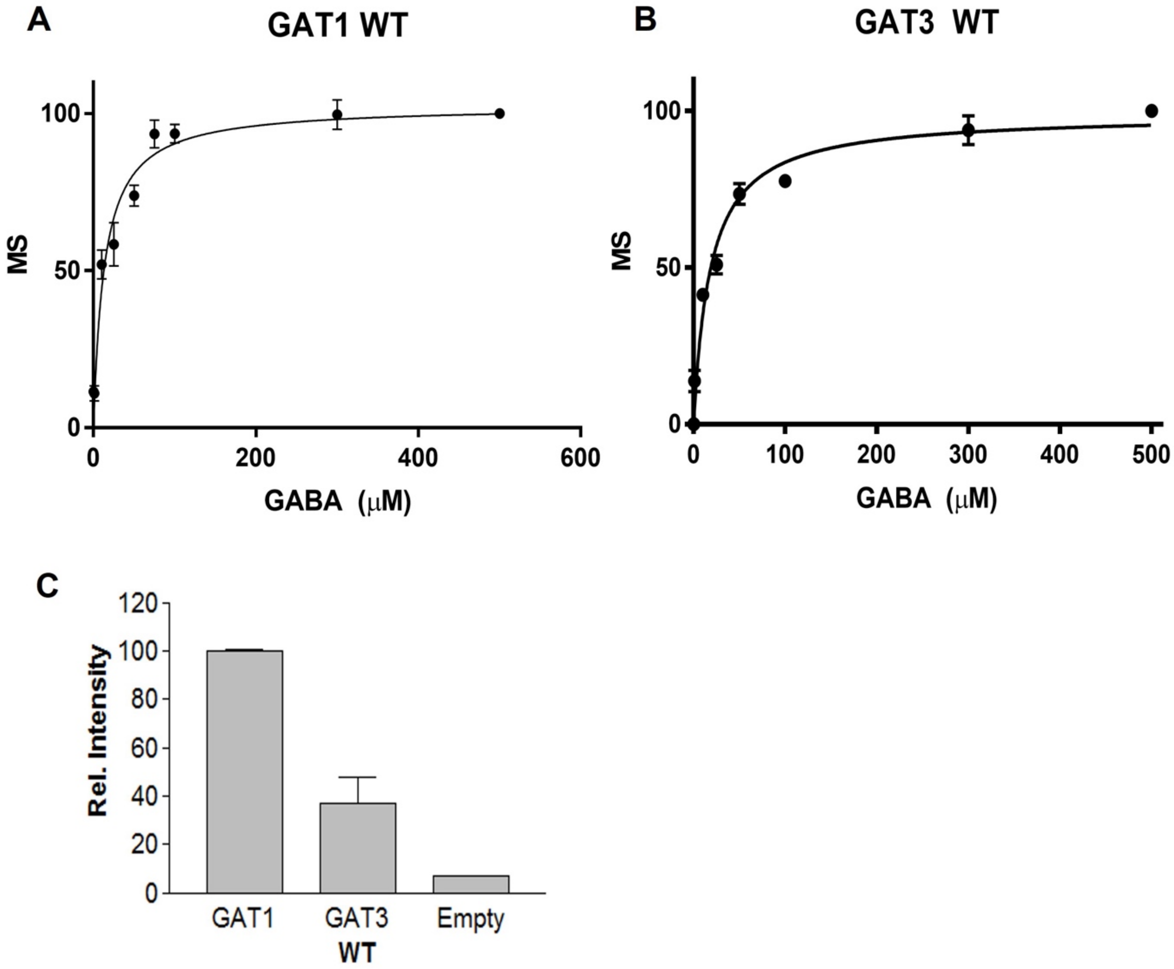


**Supplementary Figure 1:** WT dose-response curves for GAT1 and GAT3 transporters. **(A, B)** showed that GAT1 has $K_m = 13.07 \pm 2.291$ μM which is consistent with previously published GAT1 activity studies.[42] The $K_m$ of GAT3 was $18.31 \pm 2.728$ μM. **(C)** consistent with other studies the relative intensity of GAT1 uptake is faster and more specific than GAT3 (40% of relative intensity).

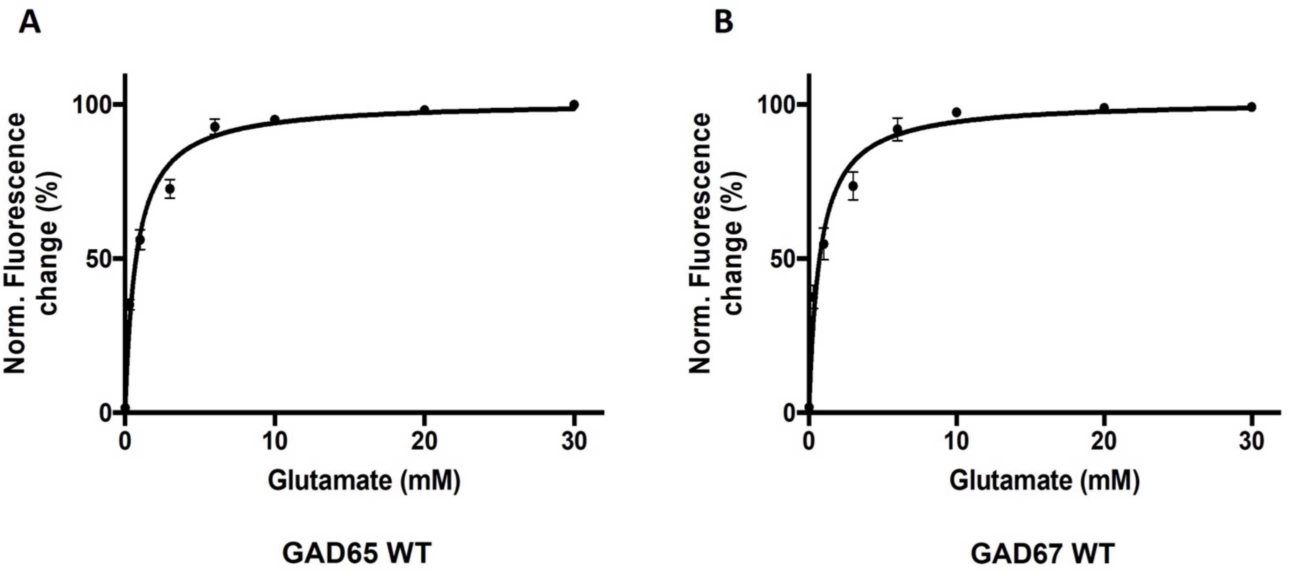


**Supplementary Figure 2:** Glutamate active dose-response curves for the enhanced GAD assay method. (**A)** GAD65 standard activity curve vs glutamate dose-response concentrations; **(B)** GAD67 standard activity curves vs glutamate dose response concentrations.

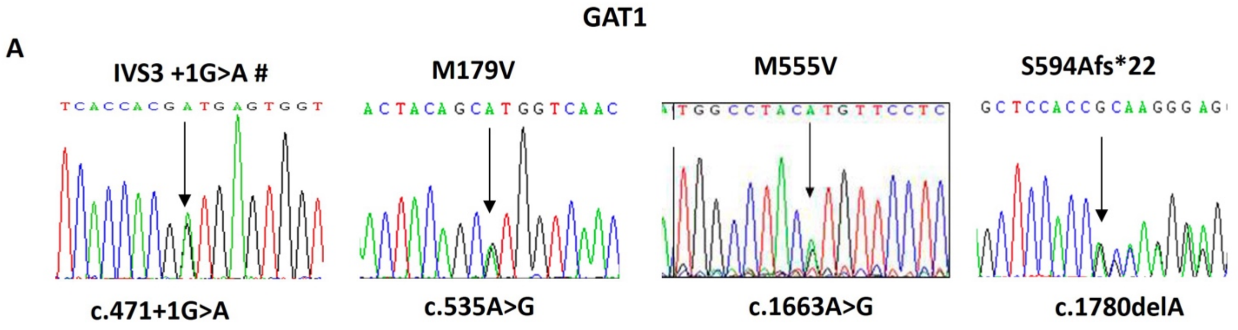


B

| ***GAT1 Variant*** | FSNYS**V**VNTT | GYMAY**V**FLTLK | SST**ARRP(+18)*** |
|---|---|---|---|
| *H.sapien (WT)* | FSNYS**M**VNTT | GYMAY**M**FLTLK | SST**S**KEAYI* |
| *P.trogdytes* | FSNYS**M**VNTT | GYMAY**M**FLTLK | SST**S**KEAYI* |
| *M.mulatta* | FSNYS**M**VNTT | GYMAY**M**FLTLK | SST**S**KEAYI* |
| *B.Trurus* | FSNYS**A**VNTT | GYMAY**M**FLTLK | GSA**S**KEAVI* |
| *C.lupus* | FSNYS**I**VNTT | GYMAY**M**FLTLK | GSA**S**KEAVI* |
| *G.gallus* | FSNYS**L**VNTT | GYMAY**M**FLTLK | NSA**N**KEAYI* |
| *R.norvegicus* | FSNYS**L**VNTT | GYMAY**M**FLTLK | SSA**S**KEAYI* |
| *M.musculus* | FSNYS**L**VNTT | GYMAY**M**FLTLK | SSA**S**KEAYI* |

**Supplementary Figure 3:** Sequence evidence in *SLC6A1* variants and GAT1 phylogenetic alignment. **(A)** Sanger sequencing traces of the *SLC6A1* genetic changes as indicated by an arrow. *SLC6A1* c.535A>G p.(Met179Val) was found in two cases but only one example of sequencing is presented (**Table 1**); # indicates further evidence seen in Figure 3 of the manuscript. **(B)** Phylogenetic alignment of the GAT1 protein changes across the species.

A

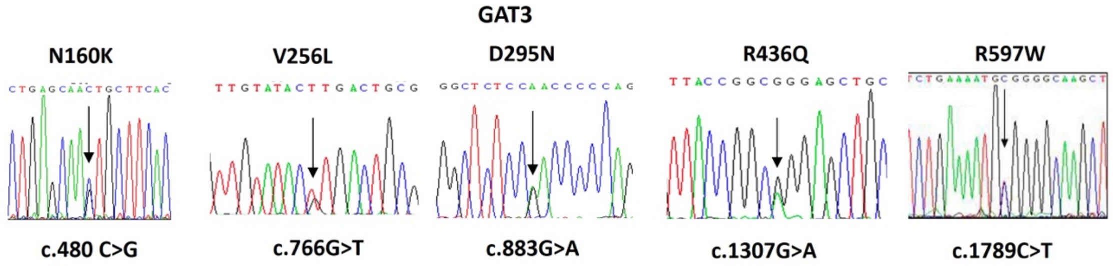


B

| | | | | | |
|---|---|---|---|---|---|
| ***GAT3 Variant*** | IFYLS**K**CFTT | GKVVY**L**TATFP | LSRLS**N**PQVW | RRGYR**Q**ELLIL | TDLKM**W**GKLG |
| *H.sapien (WT)* | IFYLS**N**CFTT | GKVVY**V**TATFP | LSRLS**D**PQVW | RRGYR**R**ELLIL | TDLKM**R**GKLG |
| *P.trogdytes* | IFYLS**N**CFTT | GKVVY**V**TATFP | LSRLS**D**PQVW | RRGYR**R**ELLIL | TDLKM**R**GKLG |
| *M.mulatta* | IFYLS**N**CFTT | GKVVY**V**TATFP | LSRLS**D**PQVW | RRGYR**R**ELLIL | TDLKM**R**GKLG |
| *B.Trurus* | IFYLS**N**CFTT | GKVVY**V**TATFP | LSRLS**D**PQVW | RRGYR**R**ELLIL | ADLKM**R**GMLG |
| *C.lupus* | IFYLS**N**CFTT | GKVVY**V**TATFP | LSRLS**D**PQVW | RRGYR**R**ELLIL | ADLKM**R**GKLG |
| *G.gallus* | IFYLS**N**CFTT | GKVVY**V**TATFP | LSRLS**D**PQVW | RRGYR**R**ELLIL | SDLKM**R**GKLG |
| *R.norvegicus* | IFYLS**N**CFTT | GKVVY**V**TATFP | LSRLS**D**PQVW | RRGYR**R**ELLIL | ADLKM**R**GKLG |
| *M.musculus* | IFYLS**N**CFTT | GKVVY**V**TATFP | LSRLS**D**PQVW | RRGYR**R**ELLIL | ADLKM**R**GKLG |

**Supplementary Figure 4:** Sequence evidence in *SLC6A11* variants and GAT3 phylogenetic alignment. **(A)** Sanger sequencing traces of the *SLC6A11* genetic changes as indicated by an arrow. **(B)** Phylogenetic alignment of the GAT3 protein changes across the species.

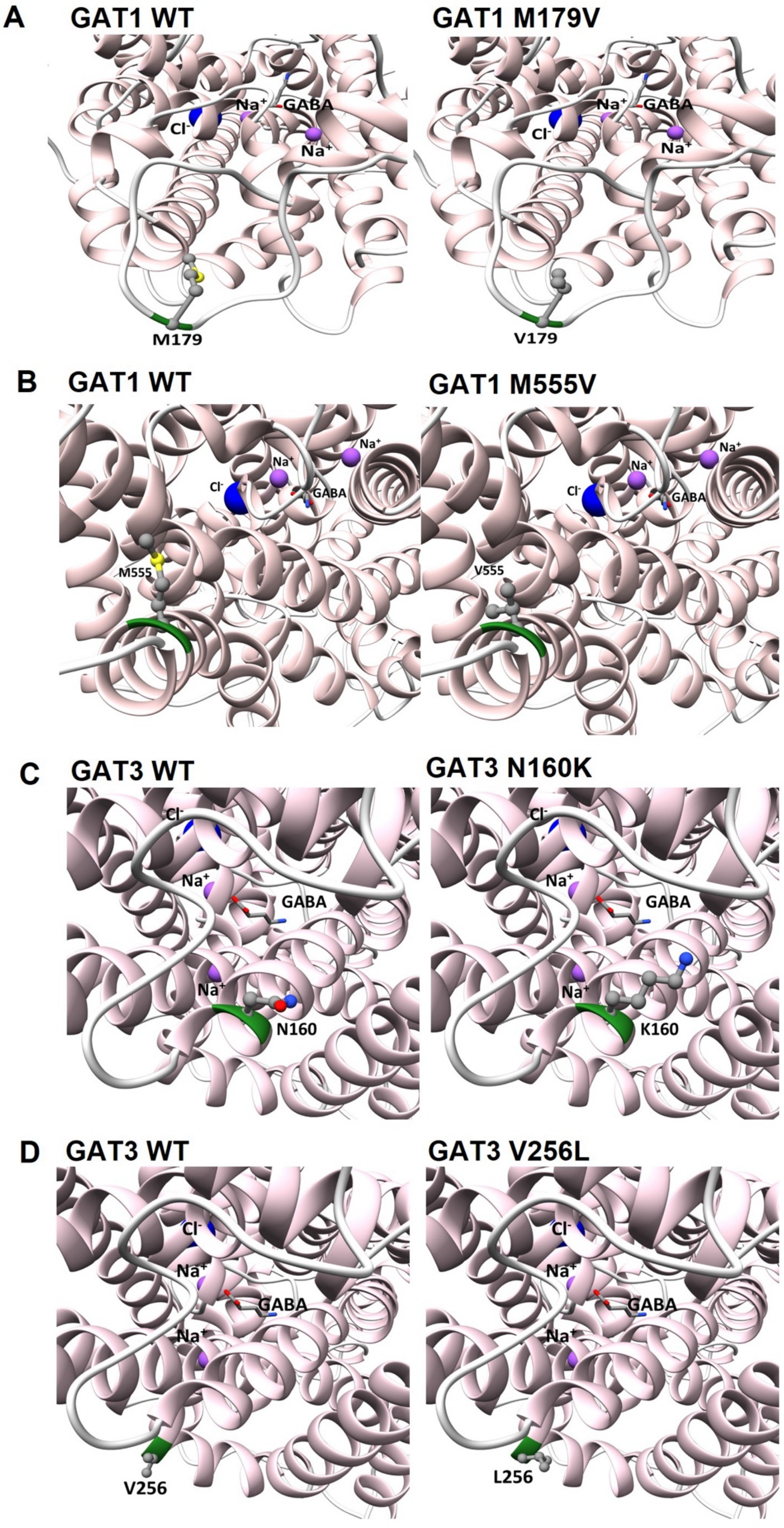
A
GAT1 WT
GAT1 M179V
Cl-
Na+
GABA
M179
V179
B
GAT1 WT
GAT1 M555V
M555
V555
C
GAT3 WT
GAT3 N160K
N160
K160
D
GAT3 WT
GAT3 V256L
V256
L256

**Supplementary Figure 5:** Molecular modelling of GABA transporter variants. **(A)** GAT1 wild type vs p.(Met179Val), showing the substitution on the extracellular loop adjacent to the entry to the transport pore; **(B)** GAT1 wild type vs p.(Met555Val), showing the substitution towards the cytoplasmic end of TM12 and potential impact on the passage of ions; **(C)** GAT3 wild type vs the p.(Asn160Lys) variant, showing the substitution at the extracellular entry to TM3 and potential for obstruction of access to the pore; **(D)** GAT1 wild type and the p.(Val256Leu) substitution towards the cytoplasmic end of TM5

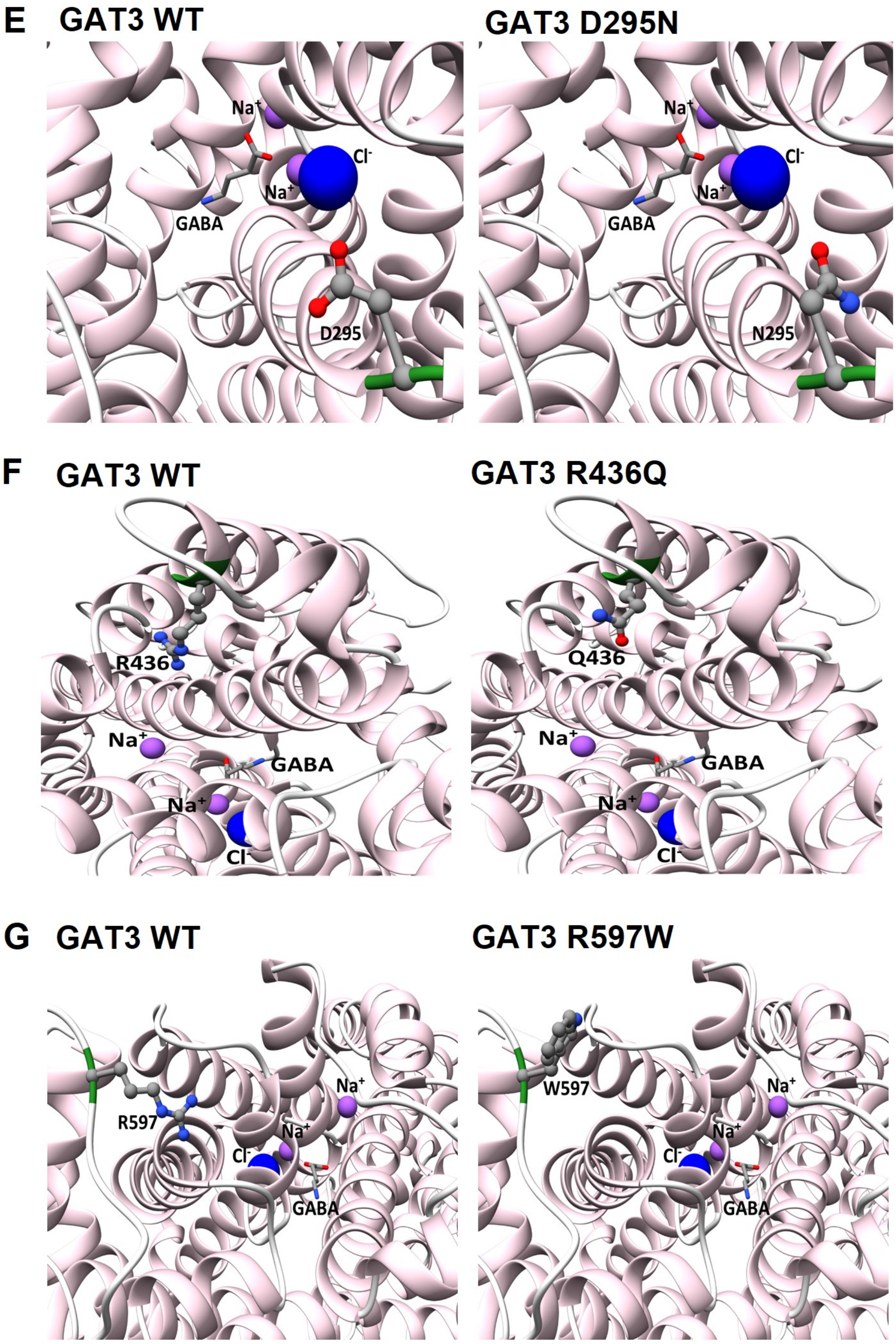


**(E)** GAT3 wild type vs p.(Asp295Asn) showing the loss of alignment of side chain groups involved in guiding the ligands and ions at the extracellular entry to the transport pore; **(F)** GAT3 wild type and the p.(Arg436Gln) substitution at the cytoplasmic end of TM9; **(G)** GAT3 WT vs p.(Arg597Trp) showing the dramatic substitution of a strong basic side chain with a bulky hydrophobic residue at the cytoplasmic exit of the pore at TM12.

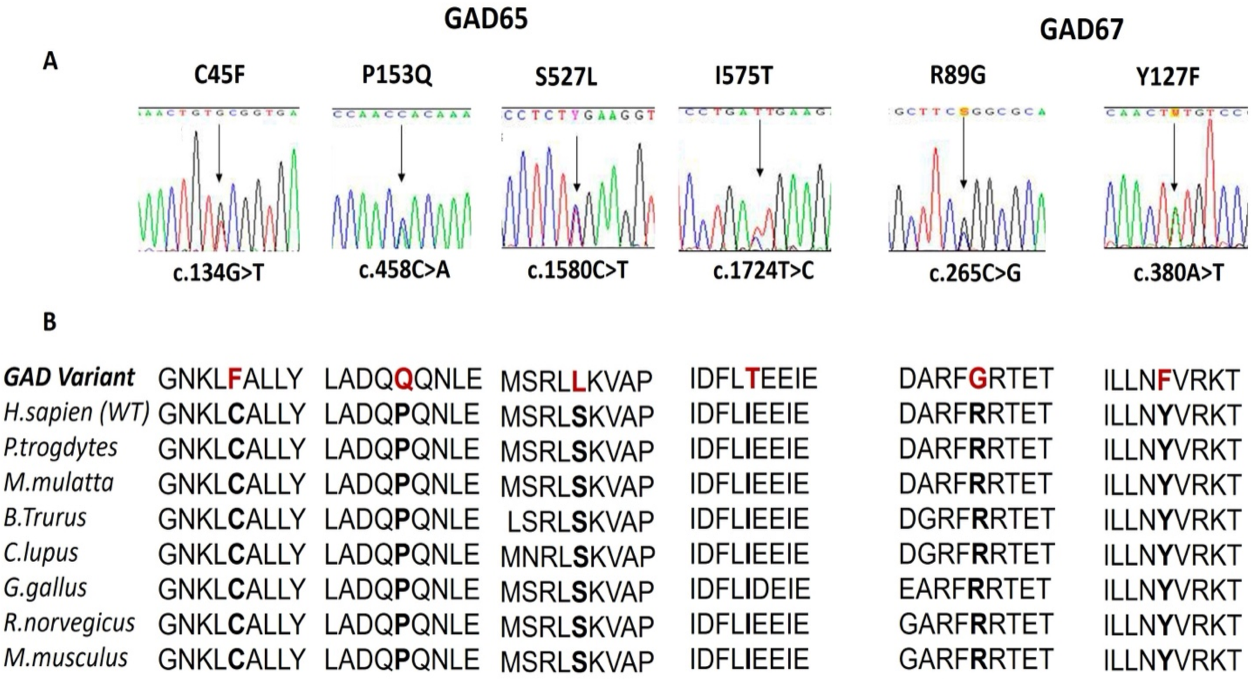


**Supplementary Figure 6:** Sequence evidence for *GAD1/GAD2* variants and GAD65/GAD67 phylogenetic alignment. **(A)** Sanger sequencing traces of the *GAD1* and *GAD2* genetic changes as indicated by an arrow. Only one representation of *GAD2* c.483C>A p.(Pro153Gln) is presented and only one representation of *GAD1* c.380A>T p.(Tyr127Phe) is presented (**Table 1**). **(B)** Phylogenetic alignment of the GAD65 and GAD67 protein changes across the species.

### Supplementary Tables

| Epilepsy Centres | Number of samples contributed to this study | Author Contributors |
|---|---|---|
| Australia | 402 | S.B., I.S., S.F.B. |
| London | 198 | K.E., R.H.T. |
| Auckland | 44 | P.S.B., M.I.R. |
| Wales | 64 | S.K.C., R.H.T. M.I.R. |

**Supplementary Table 1:** This is a breakdown of GGE, GEFS+ and Focal epilepsy cases from contributing epilepsy centres.

| Gene | Protein Variant | Case ID | cDNA Change | gnomAD MAF-Population | gnomAD MAF-European | Gene-Variant Prediction Tools *PolyPhen-2, Grantham, SIFT* |
|---|---|---|---|---|---|---|
| ***SLC6A1*** | IVS3 +1G>A | P1 | c.471+1G>A | Novel | N/A | N/D |
| ***SLC6A1*** | p.(Met179Val) | P2 / P3 | c.535A>G | 0.00154 | 0.0000339 | Benign (0.001), C15 (20.52), Tolerated (0.58) |
| ***SLC6A1*** | p.(Met555Val) | P4 | c.1663 A>G | 0.000045 | 0.0000601 | Benign (0.051), C15 (20.52), Tolerated (0.21) |
| ***SLC6A1*** | p.(Ser594Alafs*22) Frameshift | P5 | c.1780delA | Novel | N/A | N/D |
| ***SLC6A11*** | p.(Asn160Lys) | P6 | c.480C>G | 0.000001 | 0.0000008 | Possibly Damaging (0.815), C65 (93.88), Tolerated (0.25) |
| ***SLC6A11*** | p.(Val256Leu) | P7 | c.766G>T | 0.000163 | 0.000215 | Benign (0.053), C25 (83.33), Damaging (0.02) |
| ***SLC6A11*** | p.(Asp295Asn) | P8 | c.883G>A | 0.0000143 | 0.0000179 | Probably Damaging (1), C15 (23.01), Tolerated (0.12) |
| ***SLC6A11*** | p.(Arg436Gln) | P9 | c.1307G>A | 0.000097 | 0.000048 | Probably Damaging (1), C35 (42.81), Damaging (0) |
| ***SLC6A11*** | p.(Arg597Trp) | P10 | c.1789C>T | 0.000117 | 0.0000042 | Benign (0.031), C65 (101.29), Tolerated (0.08) |
| ***GAD2*** | p.(Cys45Phe) | P11 | c.134G>T | 0.0000223 | 0.0000119 | Probably Damaging (0.99), C65 (204.39), Damaging (0) |
| ***GAD2*** | p.(Pro153Gln) | P12 - P27 | c.458C>A | 0.00659 | 0.00765 | Probably Damaging (0.99), C65 (75.14), Tolerated (0.11) |
| ***GAD2*** | P(Ser527Leu) | P2 (digenic) | c.1580C>T | 0.000573 | 0.000645 | Benign (0.04), C65 (144.08), Tolerated (0.31) |
| ***GAD2*** | p.(Ile575Thr) | P28 | c.1724T>C | 0.000092 | 0.000110 | Possibly Damaging (0.76), C65 (89.28), Damaging (0.01) |
| ***GAD1*** | p.(Arg89Gly) | P29 | c.265C>G | 0.000018 | 0.0000228 | Benign (0), C65 (125.13), Tolerated (0.21) |
| ***GAD1*** | p.(Tyr127Phe) | P30 / P31 | c.380A>T | 0.000747 | 0.000948 | Possibly Damaging (0.462), C15 (21.61), Damaging (0) |

**Supplementary Table 2:** Population mean variant frequencies (MAF) were derived from 2024 v4.1.0 gnomAD data and Epi25 consortium data (NIH resources) relating to the GABAergic variants in *SLC6A1, SLC6A11, GAD1, GAD2*. However, variants selected for functional analysis were based on 2015-2016 MAF data. Genotypes are labelled under the standard genetic nomenclature guidance for molecular genetics and HGVS principles.

| Gene | Variant | HGVS Genomic Variant NC_g. | HGVS Variant Change NM_c. |
|---|---|---|---|
| ***SLC6A1*** | IVS3 +1G>A | NC_000003.12 g. | NM_ 003402.4 c.471+1G>A |
| ***SLC6A1*** | p.(Met179Val) | NC_000003.12 g.11020276A>G | NM_ 003402.4 c.535A>G |
| ***SLC6A1*** | p.(Met555Val) | NC_000003.12 g.11034666A>G | NM_ 003402.4 c.1663 A>G |
| ***SLC6A1*** | p.(Ser594Alafs*22) Frameshift | NC_000003.12 g. g.11036946delA | NM_ 003402.4 c.1780delA |
| ***SLC6A11*** | p.(Asn160Lys) | NC_000003.12 g.10819800C>G | NM_ 014229.3 c.480C>G |
| ***SLC6A11*** | p.(Val256Leu) | NC_000003.12 g.10874970G>T | NM_ 014229.3 c.766G>T |
| ***SLC6A11*** | p.(Asp295Asn) | NC_000003.12 g.10875087G>A | NM_ 014229.3 c.883G>A |
| ***SLC6A11*** | p.(Arg436Gln) | NC_000003.12 g.10929275G>A | NM_ 014229.3 c.1307G>A |
| ***SLC6A11*** | p.(Arg597Trp) | NC_000003.12 g.10938292C>T | NM_ 014229.3 c.1789C>T |
| ***GAD2*** | p.(Cys45Phe) | NC_000010.11 g.26217667G>T | NM_ 00818.3 c.134G>T |
| ***GAD2*** | p.(Pro153Gln) | NC_000010.11 g.26219214C>T | NM_ 00818.3 c.458C>A |
| ***GAD2*** | p.(Ser527Leu) | NC_000010.11 g.26292987C>T | NM_ 00818.3 c.1580C>T |
| ***GAD2*** | p.(Ile575Thr) | NC_000010.11 g.26300927T>C | NM_ 00818.3 c.1724T>C |
| ***GAD1*** | p.(Arg89Gly) | NC_000002.12 g.17905C>G | NM_ 000817.3 c.265C>G |
| ***GAD1*** | p.(Tyr127Phe) | NC_000002.12 g.19336A>T | NM_ 000817.3 c.380A>T |

**Supplementary Table 3:** Genomic reference data for the GABAergic variants using HGVS principles and data sources from NCBI dbSNP, Ensembl and Genome Data Viewer GRCh38.p14.

| Gene | cDNA Variant Change | Protein Change | Number of Cases | gnomAD MAF Population | gnomAD MAF European | Gene-Variant Prediction | Epilepsy Diagnosis | Functional Outcome |
|---|---|---|---|---|---|---|---|---|
| ***SLC6A1*** | c.1243C>A | p.(Leu415Ile) | 7 | 0.003849 | 0.004882 | Benign (0.002), CO (4.86), Tolerated (021) | CAE, JME & IPEO | Near WT Profile (not significant) **Figure 2C** |
| ***SLC6A11*** | c.424C>T | p.(His142Tyr) | 1 | 0.000129 | 0.000164 | Benign (0), C65 (83.33), Tolerated (1), CADD | JME | WT Profile **Figure 4A** |
| ***GAD2*** | c.304G>A | p.(Asp102Asn) | 1 | 0.000148 | 0.000201 | Benign (0), C15 (101.29), Tolerated (1), CADD | CAE | Near WT Profile (not significant) **Figure 6A** |
| ***GAD1*** | c.265C>G | p.(Arg89Trp) | 1 | 0.000270 | 0.0000525 | Probably Damaging (0.95), C65 (101.29), Damaging (0.03), CADD | CAE to JME | WT Profile **Figure 6B** |

**Supplementary Table 4:** Four common variants from *SLC6A1*, *SLC6A11*, *GAD1* and *GAD2* that were tested in functional assays but failed to reach significance or had parity with WT activity levels.

| Case Number | Reference |
|---|---|
| P8 | 1. Singh R *et al* (1999). Ann Neurol, 45(1): 75-81 |
| P8 | 2. Heron SE *et al* (2004). Ann Neurol, 55(4): 595-596 |
| P8 | 3. Heron SE *et al* (2007). Ann Neurol, 62(6): 560-568 |
| P11 | 4. Helbig I *et al* (2008). Lancet Neurology, 7(3): 231-245 |
| P11, P18, P30 | 5. Epi4K Consortium (2017). Lancet Neurology, 16: 135-143 |
| P14 | 6. Bagnall RD *et al* (2014). Neurology, 83(11): 1018-1021 |
| P14 | 7. Bagnall RD *et al* (2015). Ann Neurol, 79(4): 522-534 |
| P16 | 8. Winawer MR *et al* (2005). Neurology, 65(4): 523-528 |
| P16 | 9. Marini C *et al* (2004). Epilepsia, 45(5): 467-478 |
| P17 | 10. Carvill GL *et al* (2013). Nat Genet, 45(7): 825-830 |

**Supplementary Table 5:** A list of cases in manuscript **Table 1** that have featured in other studies and consortiums.
